# Chronic Shedding of a SARS-CoV-2 Alpha Variant Lineage Q.3/Q.4 in Wastewater

**DOI:** 10.1101/2023.07.26.23293191

**Authors:** Michael J. Conway, Hannah Yang, Lauren A. Revord, Avery S. Ward, Jackson D. Abel, Maggie R. Williams, Rebecca L. Uzarski, Elizabeth W. Alm

**Affiliations:** Foundational Sciences, Central Michigan University College of Medicine, Mt. Pleasant, MI; Central Michigan District Health Department, Mt. Pleasant, MI; Department of Biology, Central Michigan University; Department of Biology and Herbert H. and Grace A. Dow College of Health Professions, Central Michigan University, Mt. Pleasant MI; School of Engineering & Technology, Central Michigan University, Mt. Pleasant, MI; Institute for Great Lakes Research, Central Michigan University, Mt. Pleasant, MI

## Abstract

Central Michigan University (CMU) participated in a state-wide SARS-CoV-2 wastewater monitoring program since 2021. Wastewater samples were collected from on-campus sites and nine off-campus wastewater treatment plants servicing small metropolitan and rural communities. SARS-CoV-2 genome copies were quantified using droplet digital PCR and results were reported to the health department. One rural, off-campus site consistently produced higher concentrations of SARS-CoV-2 genome copies. Samples from this site were sequenced and initially contained predominately Alpha variant lineage Q.3, which transitioned to lineage Q.4. Alpha variant lineage Q.3/Q.4 was detected at this site beginning in fall 2021 and continued until summer 2023. Mutational analysis of reconstructed genes revealed divergence from the Alpha variant lineage Q.3 clinical sequence over time, including numerous mutations in the surface glycoprotein RBD and NTD. We discuss the possibility that a chronic SARS-CoV-2 infection accumulated adaptive mutations that promoted long-term infection. This study reveals that small wastewater treatment plants can enhance resolution of rare events and facilitate reconstruction of viral genomes due to the relative lack of contaminating sequences.

## 1. Introduction

Wastewater surveillance became an important public health tool during the COVID-19 pandemic. Wastewater surveillance programs identified outbreaks within communities and individual buildings and it is increasingly being used to detect variants of concern (Anderson-Coughlin et al., 2022; Betancourt et al., 2021; Chua et al., 2023; Corchis-Scott et al., 2021; Gibas et al., 2021; Jarvie et al., 2022; Pico-Tomàs et al., 2023; Rondeau et al., 2023; Scott et al., 2021; Vo et al., 2023; Wong et al., 2021). The goal of wastewater surveillance is to provide data to public health agencies so that they may make informed decisions regarding mitigation strategies such as physical distancing, masking, business closures, and distribution of resources such as prophylactic vaccines.

The State of Michigan Department of Health and Human Services (MDHHS) initiated a wastewater surveillance program in 2021. The program included partnerships between academic laboratories and regional public health departments that spanned large and small metropolitan areas and rural areas in both the lower and upper peninsulas. Central Michigan University (CMU) formed a partnership with the Central Michigan District Health Department (CMDHD). This partnership provided an opportunity to look at the dynamics of SARS-CoV-2 at a public university and in the surrounding small metropolitan and rural communities (Conway et al., 2023). We identified ten on-campus sewer sites and nine off-campus wastewater treatment plants (WWTPs) to sample on a weekly basis. The population sizes serviced by these WWTPs ranged from as large as 35,397 to as small as 851.

Sampling began in July 2021, which was at least seven months after emergence of the Alpha variant (B.1.1.7). The Alpha variant first appeared in North America in late November 2020 and became the predominant SARS-CoV-2 variant by the end of March 2021. The Alpha variant diverged into multiple lineages, including the Q.3 and Q.4. The Q.3 lineage is present in 5,422 clinical sequences worldwide that were uploaded to the NCBI database, and a positive sample was first collected on 7-11-20. The Q.3 lineage was detected in clinical samples from Michigan 16 times between 2-18-21 and 7-9-21. The Q.4 lineage is present in 886 clinical sequences worldwide that were uploaded to the NCBI database and was first collected on 3-15-20. The Q.4 lineage was detected in clinical samples from Michigan one time on 4-3-21.

It became clear that our smallest WWTP (estimated population served: 851) consistently produced higher concentrations of SARS-CoV-2 genome copies. Samples taken from this site from 2021-2023 were sequenced and many contained sequences that corresponded to Alpha variant lineage Q.3/Q.4. These sequences also accumulated novel mutations over time, including previously described cryptic mutations. We hypothesize that an individual was chronically infected with Alpha variant lineage Q.3 for 20-28 months. During this time, the virus adapted by accumulating novel mutations, which included previously described cryptic mutations. Importantly, we found that the earliest sample corresponded to Alpha variant lineage Q.3, which closely aligned with clinical sequences reported in summer and fall 2021; however, the sequence diverged over time and accumulated novel mutations.

These data reveal that wastewater surveillance in small metropolitan and rural communities provide an opportunity to identify novel isolates and reconstruct genes due to lower contamination with unrelated sequences. These data also suggest that humans and other animals can chronically shed SARS-CoV-2 over many months, which is associated with accumulation of adaptive mutations. Mutations associated with chronic infection may be useful to identify individuals who are chronically infected and to drive selection of appropriate therapeutics.

## 2. Materials and methods

### 2.1. Selection of sample sites

Central Michigan University (CMU) is a public research university in the City of Mt. Pleasant, Isabella County, Michigan, with an average population during the 2021-2022 academic year of 13,684 students and staff. Ten sample sites were selected on campus that collected wastewater downstream from most campus buildings, including residential halls, apartments, and academic/administrative buildings. The waste stream at these sites includes a mixture of wastewater from CMU and upstream residential areas in the City of Mt. Pleasant. Nine off-campus sites throughout the jurisdictions of the Central Michigan District Health Department (CMDHD) and Mid-Michigan District Health Department (MMDHD) were selected (Conway et al., 2023), which included the City of Mt. Pleasant, Union Township, City of Alma, City of Clare, City of Evart, three Houghton Lake townships, and Village of Marion wastewater treatment plants (WWTPs). These locations represent various land uses and population densities including urban, rural, and suburban areas, providing a large footprint of SARS CoV-2 virus shedding in Central Michigan.

### 2.2. Wastewater collection

Since July 2021, wastewater samples (500–1000 mL) were collected once each week on either Monday or Tuesday from ten sanitary sewer sites and nine WWTP influent streams (after grit removal). Sanitary sewer grab samples consisted of wastewater flowing from university dormitories and buildings and the surrounding community. Influent to WWTPs were collected as grab samples or 24-hour composite samples (Conway et al., 2023). Samples were held at 4°C no more than 48 hours before analysis.

### 2.3. Virus concentration and RNA extraction

The protocol described by Flood et al. 2021 and adopted by the Michigan wastewater surveillance network was used to concentrate virus from samples and extract viral RNA (Conway et al., 2023; Flood et al., 2021). Briefly, 100 mL wastewater or water as a negative control was mixed with 8% (w/v) molecular biology grade PEG 8000 (Promega Corporation, Madison WI) and 0.2 M NaCl (w/v). The sample was mixed slowly on a magnetic stirrer at 4 °C for 2-16 hours. Following overnight incubation, samples were centrifuged at 4,700×g for 45 min at 4 °C. The supernatant was then removed, and the pellet was resuspended in the remaining liquid, which ranged from 1-3 mL. All sample concentrates were aliquoted and stored at -80 °C until further processing. Viral RNA was extracted from concentrated wastewater using the Qiagen QIAmp Viral RNA Minikit according to the manufacturer’s protocol with previously published modifications (Qiagen, Germany) (Flood et al., 2021). In this study, a total of 200 µl of concentrate was used for RNA extraction resulting in a final elution volume of 80 µl. Extracted RNA was stored at -80 °C until analysis. A wastewater negative extraction control was included. To derive recovery efficiencies for each sample site, samples were inoculated with 10^6^ gene copies (GC)/mL Phi6 bacteriophage (Phi6) prior to the addition of PEG and NaCl. Wastewater samples were mixed, and a 1 mL sample was reserved and stored at -80 °C. RNA was extracted as stated above.

### 2.4. Detection and quantification of SARS-CoV-2

A one-step RT-ddPCR approach was used to determine the copy number/20 µL of SARS-CoV-2, and data were converted to copy number/100 mL wastewater for N1 and N2 targets using the method published by Flood et al., 2001. All the primers and probes used in this study were published previously (Conway et al., 2023). Droplet digital PCR was performed using Bio-Rad’s 1-Step RT-ddPCR Advanced kit with a QX200 ddPCR system (Bio-Rad, CA, USA). Each reaction contained a final concentration of 1 × Supermix (Bio-Rad, CA, USA), 20 U μL^-1^ reverse transcriptase (RT) (Bio-Rad, CA, USA), 15 mM DTT, 900 nmol l^-1^ of each primer, 250 nmol l^-1^ of each probe, 1 µL of molecular grade RNAse-free water, and 5.5 μL of template RNA for a final reaction volume of 22 μL (Conway et al., 2023; Flood et al., 2021; Lee et al., 2018; Lu et al., 2020). RT was omitted for DNA targets. Droplet generation was performed by microfluidic mixing of 20 μL of each reaction mixture with 70 μL of droplet generation oil in a droplet generator (Bio-Rad, CA, USA) resulting in a final volume of 40 μL of reaction mixture-oil emulsions containing up to 20,000 droplets with a minimum droplet count of > 9000. The resulting droplets were then transferred to a 96-well PCR plate that was heat-sealed with foil and placed into a C1000 96-deep-well thermocycler (Bio-Rad, CA, USA) for PCR amplification using the following parameters: 25 °C for 3 min, 50 °C for 1 h, 95 °C for 10 min, followed by 40 cycles of 95 °C for 30 s and 60 °C for 1 min with ramp rate of 2 °C/s 1 followed by a final cycle of 98 °C for 10 min. Following PCR thermocycling, each 96-well plate was transferred to a QX200 Droplet Reader (Bio-Rad, CA, USA) for the concentration determination through the detection of positive droplets containing each gene target by spectrophotometric detection of the fluorescent probe signal. All analyses were run in triplicate for each marker. To derive recovery efficiencies for each sample site, Phi6-spiked pre- and post-PEG concentration RNA samples were used to quantify Phi6 copy number using the previously published primers and probes (Conway et al., 2023). The degree of PCR inhibition was also quantified in each sample by spiking 10 μL of 10^5^ GC/ml Phi6 in a sample’s Buffer AVL, including positive controls that lacked wastewater.

### 2.5. Data analysis

All SARS-CoV-2 gene data were converted from GC per 20 µL reaction to GC per 100 mL wastewater sample before analysis (Conway et al., 2023; Flood et al., 2021). Non-detects (ND) were assigned their individual sample’s limit of detection for the purposes of data reporting, although any weekly on-campus or off-campus samples whose values matched the theoretical limit of detection were removed prior to statistical analysis. The limit of detection was calculated for each individual sample based on both the molecular assays’ theoretical detection limits (i.e., 3 positive droplets for RT-ddPCR; the lowest standard curve concentration for RT-qPCR) and the concentration factor of each processing method examined. All wastewater data were reported to MDHHS and uploaded to the Michigan COVID-19 Sentinel Wastewater Epidemiological Evaluation Project (SWEEP) dashboard (https://www.michigan.gov/coronavirus/stats/wastewater-surveillance/dashboard/sentinel-wastewater-epidemiology-evaluation-project-sweep).

### 2.6. Sequencing

RNA was shipped to GT Molecular (Fort Collins, CO) on dry ice. Library preparation was done using GT Molecular’s proprietary method, which utilized ARTIC 4.1 primers for SARS-CoV-2 amplicon generation (https://artic.network/ncov-2019). Amplicons were pooled and sequenced on a Miseq using 2×150bp reads. FASTQ files were analyzed using GT Molecular’s bioinformatics pipeline, and variant-calling was performed using a modified and proprietary version of Freyja (Karthikeyan et al., 2022).

### 2.7. Surface glycoprotein reconstruction and identification of novel mutations

FASTQ files from 11-9-21, 9-13-22, 4-24-23, and 5-1-23 contained reads that spanned the entire surface glycoprotein, they lacked contamination with other variants of concern based on variant calling, and they had high relative abundance of the Alpha variant lineage Q.3/Q.4. This allowed for reconstruction of a consensus surface glycoprotein gene for each of the above wastewater samples. Specifically, we uploaded a FASTA-formatted .txt files into Galaxy (https://usegalaxy.org/) that represented the SARS-CoV-2 reference surface glycoprotein gene. We then uploaded each of the paired-end FASTQ files for each wastewater sample. The Bowtie2 program was used to map reads against each reference sequence, creating individual .bam files per sample. The default setting was used for analysis. The Convert Bam program was then used to convert .bam files to FASTA multiple sequence alignments. Multiple sequence alignment files were uploaded to MEGA (https://www.megasoftware.net/) and converted to amino acid sequence The consensus amino acid sequence from each of these samples was manually reconstructed and then aligned with the SARS-CoV-2 surface glycoprotein reference sequence and a consensus Alpha variant lineage Q.3 sequence derived from 16 clinical samples collected in Michigan from 2-18-21 to 7-9-21. The Q.3 lineage was chosen because the earliest wastewater sample that tested positive for SARS-CoV-2 Alpha variant (i.e., 10-26-21) was Alpha variant lineage Q.3 based on the variant calling pipeline. Mutations that were present in wastewater samples but not in the SARS-CoV-2 surface glycoprotein reference sequence or clinical sample were characterized as novel mutations. FastQC was used to quantify the total number of reads in each FASTQ file, the total number of reads that aligned to the reference surface glycoprotein, the read length, and the number of poor-quality sequences (Supplementary Table 1).

### 2.8 Novel mutation and cryptic sequence hotspot analyses

We identified novel mutations as described above. Previous literature also identified cryptic sequence hotspots in SARS-CoV-2 surface glycoprotein (Gregory et al., 2022; Smyth et al., 2022). We tracked the percent prevalence of novel and cryptic sequences in wastewater samples that were positive for Alpha variant lineage Q.3/Q.4. Specifically, we uploaded FASTA-formatted .txt files into Galaxy (https://usegalaxy.org/) that represented the SARS-CoV-2 reference surface glycoprotein. We then uploaded each of the paired-end FASTQ files for each wastewater sample. The Bowtie2 program was used to map reads against the reference sequence. The default setting was used for analysis. The Convert Bam program was then used to convert .bam files to FASTA multiple sequence alignments. Multiple sequence alignment files were uploaded to MEGA (https://www.megasoftware.net/) and converted to amino acid sequence for open-reading frame analysis. Novel and cryptic sequences were identified manually, and the column of reads were copied and pasted into Excel. The column was selected, and the Analyze Data tool was selected to calculate the percent prevalence of the novel and cryptic sequences. This was repeated for each novel and cryptic sequence across all samples positive for Alpha variant lineage Q.3/Q.4 and the percent prevalence data was represented as heatmaps. Novel mutations present in the 2021, 2022, and 2023 consensus surface glycoprotein were mapped onto the furin cleaved spike protein of SARS-CoV-2 with one RBD erect using UCSF Chimera (Wrobel et al., 2020). This atomic structure was selected because it had the greatest resolution of each amino acids across the surface glycoprotein and allowed mapping of most novel mutations.

## 3. Results

### 3.1. Chronic shedding of Alpha variant lineage Q.3/Q.4 at a rural WWTP

Wastewater samples were collected between July 2021 and June 2023 from ten on-campus sanitary sewer sites and nine WWTP influent streams. SARS-CoV-2 genome copies per 100 mL wastewater were determined each week and reported to MDHHS. One site was notable for higher peaks of virus shedding, which culminated in a peak that was 4 logs higher than the mean for all sites, although high peaks of activity were observed since 9-21-21 (Figure 1). In order to identify the SARS-CoV-2 variant responsible for this activity, RNA extracted from stored wastewater concentrates was shipped to GT Molecular and their NGS and variant calling pipeline was used. RNA from the site of interest and neighboring sites were analyzed as a control. The site of interest contained high relative abundance of Delta variant lineage AY.25.1 at the first time point tested (i.e., 9-21-21) (Figure 1, Table 1). This corresponded to the beginning of the Delta variant wave in Central Michigan (Conway et al., 2023). The site of interest began shedding Alpha variant lineage Q.3/Q.4 during the next two time points tested (i.e., 10-26-21 and 11-9-21) (Figure 1, Table 1). This was preceded by sequencing data from clinical samples, which revealed 16 Alpha variant lineage Q.3 isolates collected from 2-18-21 to 7-9-21 and 1 Alpha variant lineage Q.4 isolated on 4-3-21 (Table 2). The site of interest had high relative abundance of Omicron variant lineages during the next two time points tested (i.e., 3-14-22 and 4-25-22) (Figure 1, Table 1). This corresponded to the end of the first Omicron wave in Central Michigan (Conway et al., 2023). Alpha variant lineage Q.4 became the dominant isolate in all remaining wastewater samples from the site of interest in all 2022 and 2023 samples tested, with relative abundance ranging from 47.1-98.0%. Alpha variant lineage Q.4 was also detected in the closest neighboring WWTP on 4-10-23, which corresponded to a large peak in virus shedding at that site (Figure 1, Table 1). Other sites contained Omicron variant lineages BG.5, XBB.1.5, XBB.1.5.23, XBB.1.28, XBB.1.5.1, XBB.1.5.17, XBB.1.5.49, and Delta variant lineage DT.2 at varying relative abundance (Table 1).

**Figure 1.**
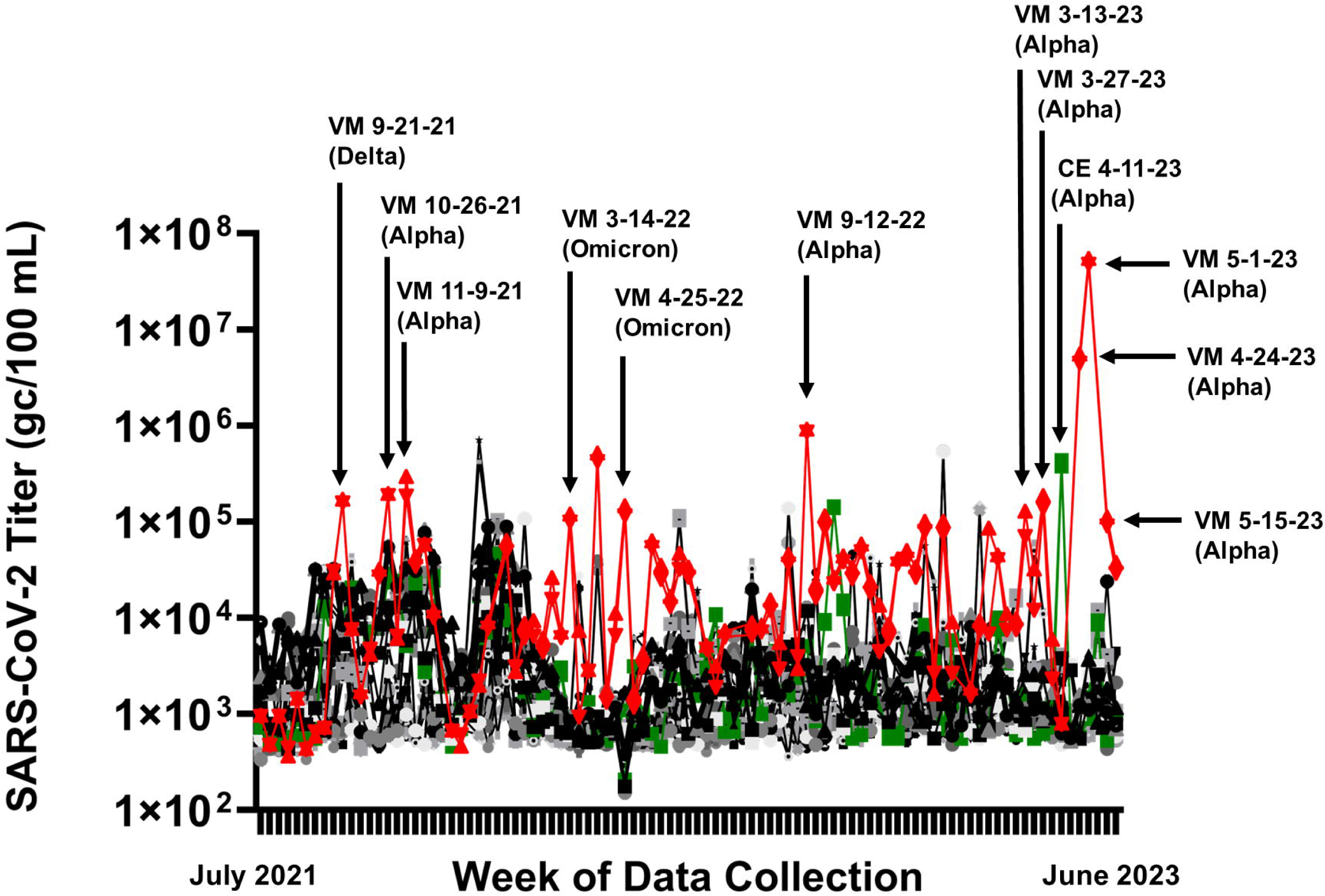
SARS-CoV-2 genome copies (GC)/100 mL wastewater detected at each weekly sample site from July 2021 to June 2023. Two letter site codes and dates are shown that correspond to sequenced samples and the variant that was identified in the highest relative abundance is indicated in parentheses.

**Table 1.** GT Molecular Variant Calling.

| Location code | Sample date | VOC (%)<sup>a</sup> | Lineage(s) (%)<sup>b</sup> |
| --- | --- | --- | --- |
| VM | 9-21-21 | Delta (98.4) | AY.25.1 (84.8) |
| VM | 10-26-21 | Alpha (94.2) | Q.3 (94.2) |
| VM | 11-9-21 | Alpha (94.9) | Q.4 (94.7) |
| VM | 3-14-22 | Omicron (93.8) | XBB.2.3.11 (74.3) |
| VM | 4-25-22 | Omicron (95.1) | XBB.1.5.81 (17.7) |
|  |  |  | XBB.1.16.1 (17.5) |
|  |  |  | XBB.1.5.10 (15.0) |
|  |  |  | EG.5 (7.7) |
|  |  |  | XBB.1.5.72 (7.4) |
|  |  |  | GJ.1.2 (5.9) |
|  |  |  | FD.1.1 (5.1) |
| VM | 9-12-22 | Alpha (97.3) | Q.4 (96.9) |
| VM | 3-13-23 | Alpha (98.0) | Q.4 (65.5) |
| VM | 3-28-23 | Omicron (27.1) | Q.4 (47.1) |
|  |  | Alpha (47.1) | B.1.1.28 (16.2) |
|  |  |  | XBB.1.5.49 (7.9) |
| VM | 4-24-23 | Alpha (97.9) | Q.4 (89.7) |
| VM | 5-1-23 | Alpha (97.4) | Q.4 (97.1) |
| VM | 5-15-23 | Alpha (96.6) | Q.4 (93.8) |
| C6 | 1-2-22 | Omicron (89.9) | BA.1.1 (82.5) |
|  |  | Delta (7.6) | B.1.617.2 (7.3) |
| CA | 6-6-22 | Omicron (99.8) | BG.5 (99.8) |
| MP | 2-20-23 | Omicron (98.4) | XBB.1.5 (45.1) |
| CL | 2-28-23 | Omicron (99.1) | DT.2 (49.9) |
|  |  |  | BQ.1.1 (16.1) |
|  |  |  | BQ.1.1.37 (16.1) |
|  |  |  | BQ.1.1.52 (16.1) |
| UT | 4-3-23 | Omicron (86.5) | XBB.1.5 (19.6) |
|  |  | Delta (7.4) | XBB.1.5.23 (15.9) |
|  |  |  | XBB.1.28 (15.6) |
|  |  |  | XBB.1.5.1 (8.2) |
|  |  |  | XBB.1.5.17 (7.5) |
| CE | 4-10-23 | Alpha (89.0) | Q.4 (88.8) |

*a, Relative abundance of variants of concern (VOC) as a percentage*
*b, Relative abundance of VOC lineages as a percentage*

**Table 2.** SARS-CoV-2 Alpha Variant Q.3/Q.4 Clinical Sequence from Michigan.

| Accession | Organism | Pangolin | Geo location | Host | Isolation source | Collection date |
| --- | --- | --- | --- | --- | --- | --- |
| OL892482 | SARS-CoV-2 | Q.3 | USA: Michigan | Homo sapiens | swab | 2/18/2021 |
| MZ158978 | SARS-CoV-2 | Q.3 | USA: Michigan | Homo sapiens | unknown | 3/29/2021 |
| MW991267 | SARS-CoV-2 | Q.3 | USA: Michigan | Homo sapiens | unknown | 3/31/2021 |
| MZ025257 | SARS-CoV-2 | Q.3 | USA: Michigan | Homo sapiens | unknown | 4/4/2021 |
| OL812274 | SARS-CoV-2 | Q.3 | USA: Michigan | Homo sapiens | swab | 4/8/2021 |
| MZ071109 | SARS-CoV-2 | Q.3 | USA: Michigan | Homo sapiens | oronasopharynx | 4/12/2021 |
| MZ131434 | SARS-CoV-2 | Q.3 | USA: Michigan | Homo sapiens | oronasopharynx | 4/16/2021 |
| OL803302 | SARS-CoV-2 | Q.3 | USA: Michigan | Homo sapiens | swab | 4/23/2021 |
| MZ416155 | SARS-CoV-2 | Q.3 | USA: Michigan | Homo sapiens | oronasopharynx | 5/12/2021 |
| MZ353922 | SARS-CoV-2 | Q.3 | USA: Michigan | Homo sapiens | oronasopharynx | 5/20/2021 |
| OK296641 | SARS-CoV-2 | Q.3 | USA: Michigan | Homo sapiens | unknown | 5/24/2021 |
| OK233076 | SARS-CoV-2 | Q.3 | USA: Michigan | Homo sapiens | oronasopharynx | 5/25/2021 |
| OK292910 | SARS-CoV-2 | Q.3 | USA: Michigan | Homo sapiens | unknown | 6/12/2021 |
| OK187847 | SARS-CoV-2 | Q.3 | USA: Michigan | Homo sapiens | unknown | 6/18/2021 |
| MZ780743 | SARS-CoV-2 | Q.3 | USA: Michigan | Homo sapiens | unknown | 6/18/2021 |
| MZ728995 | SARS-CoV-2 | Q.3 | USA: Michigan | Homo sapiens | unknown | 7/9/2021 |
| MZ025226 | SARS-CoV-2 | Q.4 | USA: Michigan | Homo sapiens | unknown | 4/3/2021 |

### 3.2. Accumulation of novel mutations in the RBD and NTD

We reasoned that chronic shedding of Alpha variant lineage Q.3/Q.4 would lead to accumulation of novel or cryptic mutations that do not align with sequences identified in most clinical and wastewater samples. Alignment of reconstructed consensus genes with the SARS-CoV-2 surface reference gene and a consensus Alpha variant lineage Q.3 clinical sequence revealed that the surface glycoprotein harbored 9 novel mutations in 2021, 25 novel mutations in 2022, and 38 novel mutations in 2023 (Supplemental Figure 1). We expanded this analysis by quantifying the percent prevalence of each of the 38 novel mutations identified in the 2023 samples across all wastewater samples that were positive for Alpha variant lineage Q.3/Q.4. A heatmap showed that these mutations accumulated within the population over time, while also retaining diversity at each position (Figure 2). This analysis was also performed using previously published cryptic mutations (Gregory et al., 2022; Smyth et al., 2022). The cryptic mutations that were selected over time included N460K, L828F, E484A, and Y505H.

**Figure 2.**
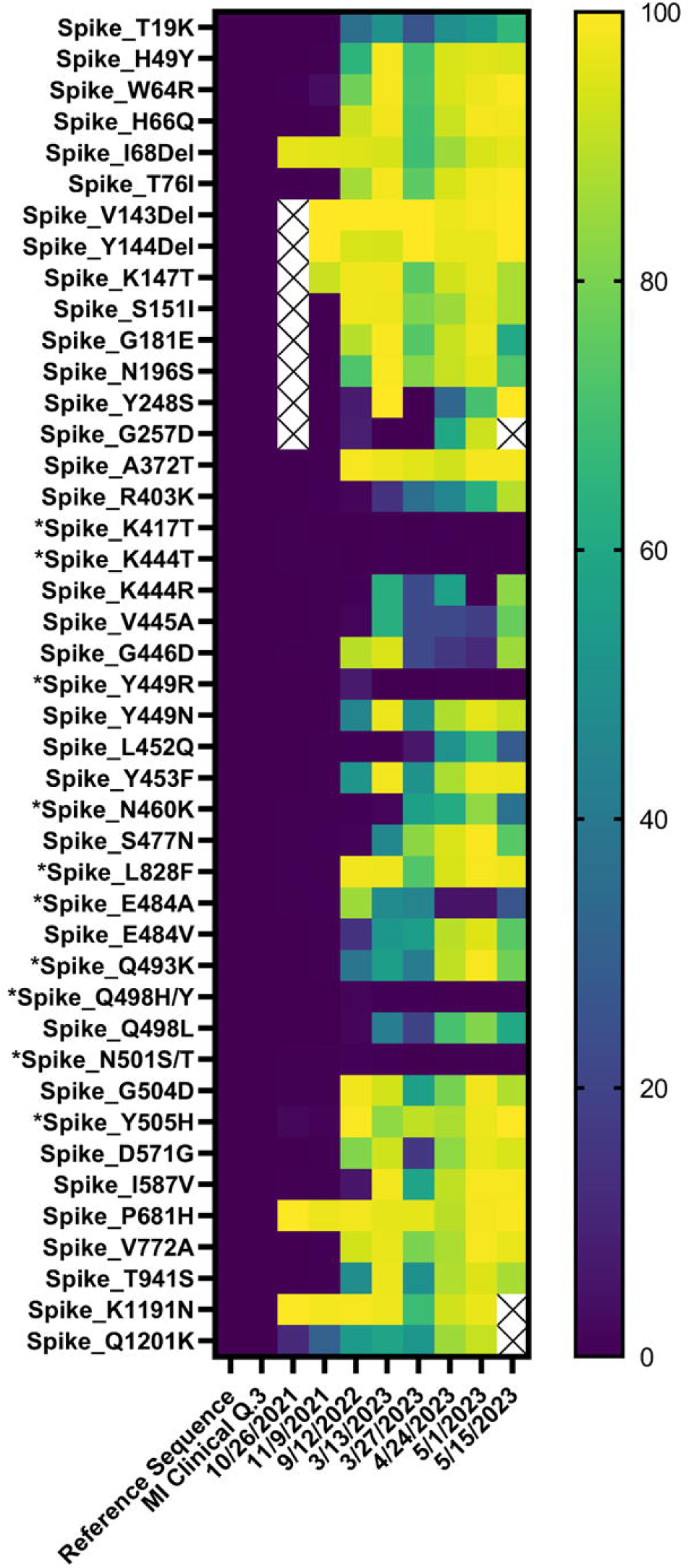
Heatmap showing the percent prevalence of novel and previously identified cryptic mutations (*) in each wastewater sample that was positive for Alpha variant lineage Q.3/Q.4 (Gregory et al., 2022; Smyth et al., 2022). Empty cells represent mutations that had fewer than 3 reads.

The majority of the novel mutations accumulated in the receptor binding domain (RBD) and N-terminal domain (NTD) of the surface (a.k.a., spike) glycoprotein (Figure 3A-F). The following mutations were identified in the RBD: A372T, R403K, K444R, V445A, G446D, Y449N, L452Q, Y453F, N460K, S477N, E484V, Q493K, Q498L, G504D, and Y505H. The following mutations were identified in the NTD: T19K, H49Y, W64R, H66Q, I68Del, T76I, V143Del, Y144Del, K147T, S151I, G181E, N196S, Y248S, and G257D. Additional mutations were identified toward the C-terminus: D571G, I587V, P681H, V772A, L828F, T941S, V1176F, K1191N, and Q1201K. Many of these mutations were not mapped onto the surface glycoprotein due to a lack of resolution toward the C-terminus.

**Figure 3.**
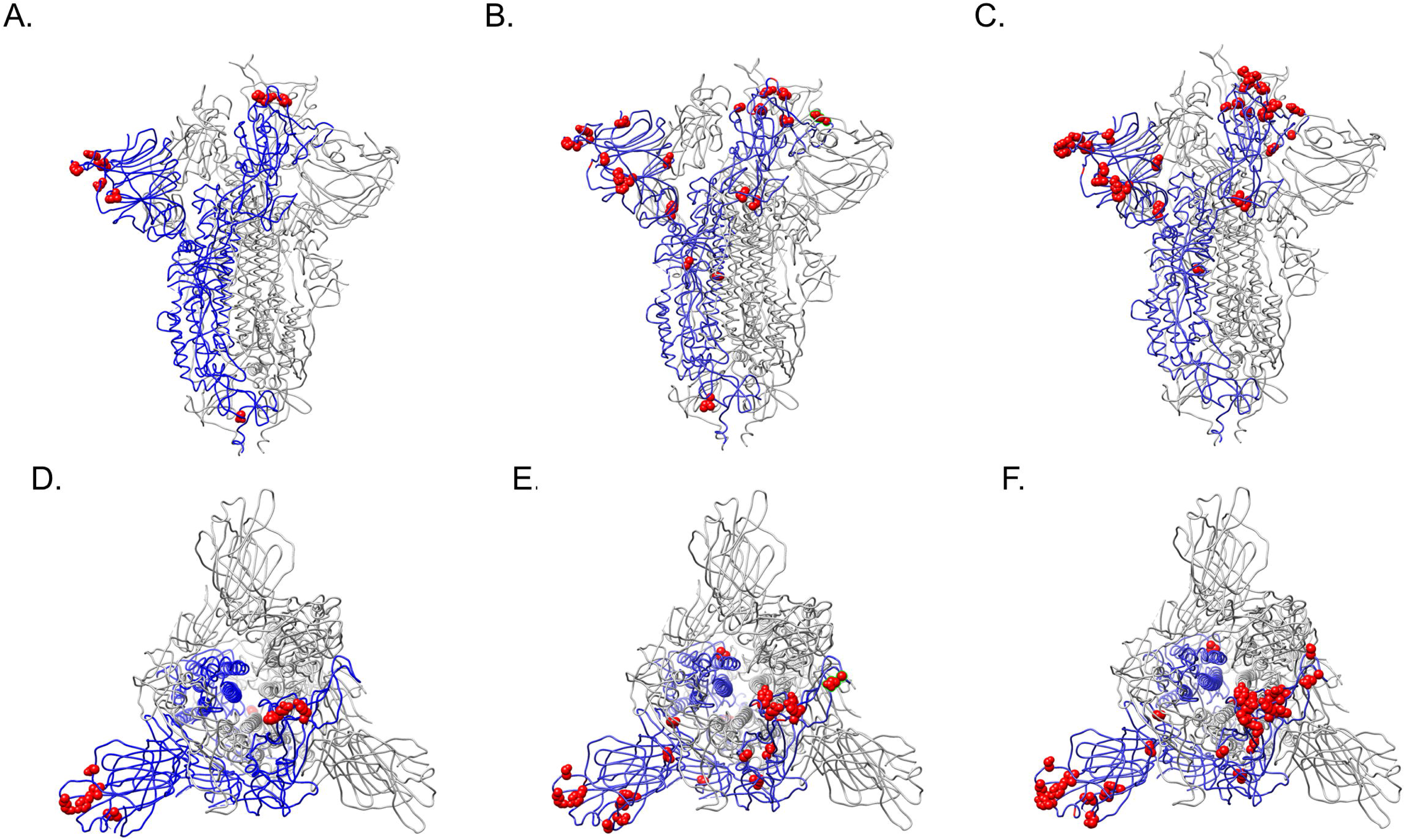
Novel mutations present in the (A, D) 2021, (B, E) 2022, and (C, F) 2023 consensus surface glycoproteins were mapped onto the furin cleaved spike protein of SARS-CoV-2 with one RBD erect represented as red Corey-Pauling_Koltun (CPK) spheres using UCSF Chimera (Wrobel et al., 2020).

The closest neighboring WWTP also contained Alpha variant lineage Q.4 (Table 1). Alignment of the reconstructed surface glycoprotein from CE 4-10-23 revealed shared mutations with reconstructed surface glycoproteins from 11-9-21, 9-12-22, and 5-1-23, and four unique mutations: L24S, H245Y, G446A, and Y1155F (Supplementary Figure 2). The mutations shared with the reconstructed surface glycoproteins from 11-9-21, 9-22-22, and 5-1-23 suggested that CE 4-10-23 shared a common ancestor with more recent isolates.

## 4. Discussion

Central Michigan University participated in a statewide SARS-CoV-2 wastewater surveillance network starting July 2021. Two major waves of SARS-CoV-2 passed through the central Michigan region during the 2021/2022 academic year, which were characterized by the emergence of Delta and Omicron variants (Conway et al., 2023). Delta and Omicron variants were preceded by the Alpha variant B.1.1.7. The SARS-CoV-2 Alpha variant B.1.1.7 wave passed through central Michigan during winter/spring 2021. Distinct Alpha variant subpopulations were also present at that time including Q.3 and Q.4 lineages.

Retrospective analysis of wastewater data revealed that one rural site produced consistently higher concentrations of SARS-CoV-2 copy numbers since September 2021. NGS sequencing revealed that this site began shedding Alpha variant lineage Q.3 by October 2021 and that this continued to at least May 2023. Clinical sequence data revealed that Alpha variant lineage Q.3/Q.4 was present in Michigan between February to July 2021. This preceded the start of wastewater surveillance in central Michigan and our first detection of Alpha variant lineage Q.3/Q.4 in wastewater by 3-8 months. It is unclear how many individuals were originally infected with this lineage at the site of interest, and it is unclear how many individuals continued to shed the virus into the sewer shed. However, due to the small population served at this rural WWTP, our November 2021 Alpha variant lineage Q.4 surface glycoprotein reconstruction likely represents a chronic infection that lasted for 3-8 months. At this stage of the chronic infection, Alpha variant lineage Q.4 already accumulated 9 novel mutations in the surface glycoprotein.

Most of the mutations reside in the surface glycoprotein RBD and NTD. These domains are critical for host receptor binding and also contain key epitopes leveraged by the adaptive immune system to control and prevent repeat infection. A striking mutation that developed in 2023 was R403K. This converted the RGD receptor binding motif to KGD, which is present in SARS-CoV-1 – a historically more lethal yet less transmissible virus (Luan et al., 2020; Makowski et al., 2021; Norris et al., 2023). This is particularly interesting since R403 is highly conserved in SARS-CoV-2 surface glycoproteins and only 294 of ∼3.4 million sequences recorded on GSAID contained a conservative change of R403K (Zech et al., 2021). Many other mutations have also been previously characterized. For instance, engineering the A372T mutation into SARS-CoV-2 reduced binding to ACE2 and enhanced replication in human lung cells (Kang et al., 2021). K444R, V445A, G446D, Y449N, L452Q, N460K, S477N, and E484V (and cryptic mutation E484A) have been associated with resistance to antibody-mediated neutralization, and N460K was previously observed during a persistent infection in an immunocompromised patient (Greaney et al., 2021a; Greaney et al., 2021b; Gregory et al., 2022; Kimura et al., 2022; Ku et al., 2021; Liu et al., 2021; Tortorici et al., 2020; VanBlargan et al., 2021; Wang et al., 2022; Wang et al., 2023). E848V also reduced ACE2 binding (Greaney et al., 2021b). L452Q had higher binding to soluble ACE2 (Kimura et al., 2022). Y453F increased binding to mink ACE2 (Ren et al., 2021). Q493K increased binding to mouse ACE2 but was also present in the Omicron variant, suggesting that is promoted immune evasion and modified receptor binding. It also developed in an immunocompromised patient undergoing convalescent plasma treatment (Chen et al., 2021; Huang et al., 2021; Zhang et al., 2021). Q498L was predicted to lower stability of surface glycoprotein and ACE2 interaction but no studies are available to confirm this prediction (Mishra et al., 2022). G504D is associated with immune evasion; however, the G504D substitution is rarely observed in SARS-CoV-2 strains, with a mutant rate below 0.002% (Chen et al., 2023; VanBlargan et al., 2021). Y505H was in all lineages of the Omicron variant suggesting that it enhanced immune evasion and receptor binding (Zhou et al., 2023).

In the NTD, H49Y impacts surface glycoprotein structure and influences binding of several antiviral compounds and increased resistance to vaccine sera (Sixto-López et al., 2021; Wang et al., 2023). T76I increases infectivity in the Lambda variant, although it is suggested that it behaves as a compensatory mutation (Kimura et al., 2022). T76I also effects antibody binding and immune escape (Ou et al., 2022). V143Del is present in Omicron variants suggesting that it enhanced immune evasion and receptor binding (Cao et al., 2022). Y144Del may play a role in ACE2 receptor binding or neutralizing antibody escape and deletions in this region were identified in immunocompromised patients (Chen et al., 2021; Goes et al., 2022; Shen et al., 2021). T19K, W64R, H66Q, I68Del, K147T, S151I, G181E, N196S, Y248S, and G257D substitutions have not been characterized.

Toward the C-terminus, P681H leads to escape of IFITM restriction and is necessary for type I interferon resistance (Lista et al., 2022). L828F is a highly prevalent cryptic mutation, which has an unknown origin, although likely due to shedding from chronically infected humans or animals (Gregory et al., 2022; Smyth et al., 2022). D571G, I587V, V772A, T941S, V1176F, K1191N, and Q1201K substitutions have not been characterized.

Collectively, the mutations that have accumulated in the surface glycoprotein gene are likely a response to the host’s innate and adaptive immune systems, and perhaps due to long-term persistence in an immunosuppressed patient and adaptation to any prophylactic or targeted drugs used to clear the infection (Ahmadi et al., 2023; Chen et al., 2021; Goes et al., 2022; Kemp et al., 2021). The presence of previously identified cryptic mutations suggest that these mutations may predict a chronic infection. It is important to note that the site of interest is not directly discussed in this manuscript in order to retain the anonymity of the person(s) responsible for chronically shedding this virus. However, we would like to highlight the potential of wastewater surveillance and possibly fecal testing for identification of long COVID. This would be particularly useful if convergent mutations emerge during a chronic infection that are predictive of this condition. The spectrum of mutations might guide appropriate selection of antivirals and antibody-based therapies. Additional mutations outside of the surface glycoprotein gene were also present in these samples but not analyzed for this manuscript. It is likely that mutations outside of the surface glycoprotein gene are also important to facilitate chronic infection.

In summary, these data support that an individual can be chronically infected with SARS-CoV-2 over many months and possibly a few years. During this time, SARS-CoV-2 can accumulate many mutations in the surface glycoprotein, which concentrate in the RBD and NTD. Further research is needed to determine if these mutations are predictive of chronic infection and if they can be used as a biomarker in individuals with Long COVID and leveraged to tailor selection or development of pharmaceutical therapies. Additionally, this study shows that small WWTPs can enhance the resolution of rare biological events and allow for total reconstruction of viral genes and their corresponding proteins.

## Data Availability

All data produced in the present study are available upon reasonable request to the authors

## Acknowledgements

We thank MDHHS and MiNET for supporting wastewater collection, processing, and data analysis. We also thank the WWTP staff who provided samples every week during a pandemic. We thank CMU undergraduate assistants Justus Holben, Gabrielle Reau, Jessica Broach, Hamzah Khan, Jayde-Ann Taylor, Ashley Bergmooser, Kaitlyn Perry, Emily Rosema, and Alexis Bruce for their support. This is contribution number XXX of the Central Michigan University Institute for Great Lakes Research.

---

**Supplemental Figure 1. Clustal Omega alignment of reconstructed 2021, 2022, and 2023 surface glycoproteins with reference and consensus Alpha variant Q.3 clinical sequence surface glycoprotein.**

**Supplemental Figure 2. Clustal Omega alignment of reconstructed 2021, 2022, and 2023 surface glycoproteins with reconstructed glycoprotein from CE 4-10-23. Mutations unique to CE-4-10-23 are highlighted in grey.**

**Supplemental Table 1. MEGA and FastQC analyses of wastewater samples positive for Alpha variant lineage Q.3/Q.4**

