## Supplemental Table 1 for "Chronic Shedding of a SARS-CoV-2 Alpha Variant Lineage Q.3/Q.4 in Wastewater"

Supplemental Table 1. MEGA and FastQC analyses of wastewater samples positive for Alpha variant lineage Q.3/Q.4

| Sample | Spike reads^a^ | Read length^b^ | Poor quality sequences^c^ |
| --- | --- | --- | --- |
| VM 10-26-21 | 115,146 | 151 | 0 |
| VM 11-9-21 | 154,428 | 151 | 0 |
| VM 9-12-22 | 132,132 | 151 | 0 |
| VM 3-13-23 | 189,826 | 151 | 0 |
| VM 3-27-23 | 84,406 | 151 | 0 |
| VM 4-24-23 | 156,360 | 151 | 0 |
| VM 5-1-23 | 229,314 | 151 | 0 |
| VM 5-15-23 | 37,054 | 151 | 0 |
| CE 4-10-23 | 29,986 | 151 | 0 |

*a, Fastq files were aligned to reference surface glycoprotein (spike) and total spike reads were identified in MEGA. b,c FastQC was used to determine read length and the number of poor quality sequences.*
