## Supplemental Figure 1 for "Chronic Shedding of a SARS-CoV-2 Alpha Variant Lineage Q.3/Q.4 in Wastewater"

```
CLUSTAL O(1.2.4) multiple sequence alignment


VM_11-9-21      MFVFLVLLPLVSSQCVNLTTRTQLPPAYTNSFTRGVYYPDKVFRSSVLHSTQDLFLPFFS	60
Reference       MFVFLVLLPLVSSQCVNLTTRTQLPPAYTNSFTRGVYYPDKVFRSSVLHSTQDLFLPFFS	60
Q.3             MFVFLVLLPLVSSQCVNLTTRTQLPPAYTNSFTRGVYYPDKVFRSSVLHSTQDLFLPFFS	60
VM-9-12-22      MFVFLVLLPLVFSQCVSLTTRTQLPPAYTNSFTRGVYYPDKVFRSSVLYSTQDLFLPFFS	60
VM_5-1-23       MFVFLVLLPLVSSQCVNLKTRTQLTPAYTNSFTRGVYYPDKVFRSSVLYSTQDLFLPFFS	60
                *********** ****.*.***** ***********************:***********

VM_11-9-21      NVTWFHA---SGTNGTKRFDNPVLPFNDGVYFASTEKSNIIRGWIFGTTLDSKTQSLLIV	117
Reference       NVTWFHAIHVSGTNGTKRFDNPVLPFNDGVYFASTEKSNIIRGWIFGTTLDSKTQSLLIV	120
Q.3             NVTWFHAI--SGTNGTKRFDNPVLPFNDGVYFASTEKSNIIRGWIFGTTLDSKTQSLLIV	118
VM-9-12-22      NVTRFQAI--SGTNGIKRFDNPVLPFNDGVYFASTEKSNIIRGWIFGTTLDSKTQSLLIV	118
VM_5-1-23       NVTRFQA---SGTNGIKRFDNPVLPFNDGVYFASTEKSNIIRGWIFGTTLDSKTQSLLIV	117
                *** *:*   ***** ********************************************

VM_11-9-21      NNATNVVIKVCEFQFCNDPFLG--YHTNNKSWMESEFRVYSSANNCTFEYVSQPFLMDLE	175
Reference       NNATNVVIKVCEFQFCNDPFLGVYYHKNNKSWMESEFRVYSSANNCTFEYVSQPFLMDLE	180
Q.3             NNATNVVIKVCEFQFCNDPFLGVY-HKNNKSWMESEFRVYSSANNCTFEYVSQPFLMHLE	177
VM-9-12-22      NNATNVVIKVCEFQFCNDPFLGV-YHTNNKIWMESEFRVYSSANNCTFEYVSQPFLMDLE	177
VM_5-1-23       NNATNVVIKVCEFQFCNDPFLG--YHTNNKIWMESEFRVYSSANNCTFEYVSQPFLMDLE	175
                **********************   *.*** **************************.**

VM_11-9-21      GKQGNFKNLREFVFKNIDGYFKIYSKHTPINLVRDLPQGFSALEPLVDLPIGINITRFQT	235
Reference       GKQGNFKNLREFVFKNIDGYFKIYSKHTPINLVRDLPQGFSALEPLVDLPIGINITRFQT	240
Q.3             GKQGNFKNLREFVFKNIDGYFKIYSKHTPINLVRDLPQGFSALEPLVDLPIGINITRFQT	237
VM-9-12-22      EKQGNFKNLREFVFKSIDGYFKIYSKHTPINLVRDLPQGFSALEPLVDLPIGINITRFQT	237
VM_5-1-23       EKQGNFKNLREFVFKSIDGYFKIYSKHTPINLVRDLPQGFSALEPLVDLPIGINITRFQT	235
                 **************.********************************************

VM_11-9-21      LLALHRSYLTPGDSSSGWTAGAAAYYVGYLQPRTFLLKYNENGTITDAVDCALDPLSETK	295
Reference       LLALHRSYLTPGDSSSGWTAGAAAYYVGYLQPRTFLLKYNENGTITDAVDCALDPLSETK	300
Q.3             LLALHRSYLTPGDSSSGWTAGAAAYYVGYLQPRTFLLKYNENGTITDAVDCALDPLSETK	297
VM-9-12-22      LLALHRSYLTPGDSSSGWTAGAAAYYVGYLQPRTFLLKYNENGTITDAVDCALDPLSETK	297
VM_5-1-23       LLALHRSSLTPGDSSSDWTAGAAAYYVGYLQPRTFLLKYNENGTITDAVDCALDPLSETK	295
                ******* ********.*******************************************

VM_11-9-21      CTLKSFTVEKGIYQTSNFRVQPTESIVRFPNITNLCPFGEVFNATRFASVYAWNRKRISN	355
Reference       CTLKSFTVEKGIYQTSNFRVQPTESIVRFPNITNLCPFGEVFNATRFASVYAWNRKRISN	360
Q.3             CTLKSFTVEKGIYQTSNFRVQPTESIVRFPNITNLCPFGEVFNATRFASVYAWNRKRISN	357
VM-9-12-22      CTLKSFTVEKGIYQTSNFRVQPTESIVRFPNITNLCPFGEVFNATRFASVYAWNRKRISN	357
VM_5-1-23       CTLKSFTVEKGIYQTSNFRVQPTESIVRFPNITNLCPFGEVFNATRFASVYAWNRKRISN	355
                ************************************************************

VM_11-9-21      CVADYSVLYNSASFSTFKCYGVSPTKLNDLCFTNVYADSFVIRGDEVRQIAPGQTGKIAD	415
Reference       CVADYSVLYNSASFSTFKCYGVSPTKLNDLCFTNVYADSFVIRGDEVRQIAPGQTGKIAD	420
Q.3             CVADYSVLYNSASFSTFKCYGVSPTKLNDLCFTNVYADSFVIRGDEVRQIAPGQTGKIAD	417
VM-9-12-22      CVADYSVLYNSTSFSTFKCYGVSPTKLNDLCFTNVYADSFVIRGDEVRQIAPGQTGKIAD	417
VM_5-1-23       CVADYSVLYNSTSFSTFKCYGVSPTKLNDLCFTNVYADSFVIKGDEVRQIAPGQTGKIAD	415
                ***********:******************************:*****************

VM_11-9-21      YNYKLPDDFTGCVIAWNSNNLDSKVGGNHNYLYRLFRKSNLKPFERDISTEIYQAGSTPC	475
Reference       YNYKLPDDFTGCVIAWNSNNLDSKVGGNYNYLYRLFRKSNLKPFERDISTEIYQAGSTPC	480
Q.3             YNYKLPDDFTGCVIAWNSNNLDSKVGGNYNYLYRLFRKSNLKPFERDISTEIYQAGSTPC	477
VM-9-12-22      YNYKLPDDFTGCVIAWNSNNLDSKVDGNNNYLFRLFRKSNLKPFERDISTEIYQAGSTPC	477
VM_5-1-23       YNYKLPDDFTGCVIAWNSNNLDS---GNNNYQFRLFRKSKLKPFERDISTEIYQAGNTPC	472
                ***********************   ** ** :******:****************.***

VM_11-9-21      NGVEGFNCYFPLQSYGFRRPTYGVGYQPYRVVVLSFELLHAPATVCGPKKSTNLVKNKCV	535
Reference       NGVEGFNCYFPLQSYGFQ-PTNGVGYQPYRVVVLSFELLHAPATVCGPKKSTNLVKNKCV	539
Q.3             NGVEGFNCYFPLQSYGFQ-PTYGVGYQPYRVVVLSFELLHAPATVCGPKKSTNLVKNKCV	536
VM-9-12-22      NGVAGFNCYFPLQSYGFR-PTYGVDHQPYRVVVLSFELLHAPATVCGPKKSTNLVKNKCV	536
VM_5-1-23       NGVVGFNCYFPLKSYGFL-PTYGVDHQPYRVVVLSFELLHAPATVCGPKKSTNLVKNKCV	531
                *** ********:****  ** **.:**********************************

VM_11-9-21      NFNFNGLTGTGVLTESNKKFLPFQQFGRDIDDTTDAVRDPQTLEILDITPCSFGGVSVIT	595
Reference       NFNFNGLTGTGVLTESNKKFLPFQQFGRDIADTTDAVRDPQTLEILDITPCSFGGVSVIT	599
Q.3             NFNFNGLTGTGVLTESNKKFLPFQQFGRDIDDTTDAVRDPQTLEILDITPCSFGGVSVIT	596
VM-9-12-22      NFNFNGLTGTGVLTESNKKFLPFQQFGRDIDGTTDAVRDPQTLEILDVTPCSFGGVSVIT	596
VM_5-1-23       NFNFNGLTGTGVLTESNKKFLPFQQFGRDIDGTTDAVRDPQTLEILDVTPCSFGGVSVIT	591
                ****************************** .***************:************

VM_11-9-21      PGTNTSNQVAVLYQGVNCTEVPVAIHADQLTPTWRVYSTGSNVFQTRAGCLIGAEHVNNS	655
Reference       PGTNTSNQVAVLYQDVNCTEVPVAIHADQLTPTWRVYSTGSNVFQTRAGCLIGAEHVNNS	659
Q.3             PGTNTSNQVAVLYQGVNCTEVPVAIHADQLTPTWRVYSTGSNVFQTRAGCLIGAEHVNNS	656
VM-9-12-22      PGTNTSNQVAVLYQGVNCTEVPVAIHADQLTPTWRVYSTGSNVFQTRAGCLIGAEHVNNS	656
VM_5-1-23       PGTNTSNQVAVLYQGVNCTEVPVAIHADQLTPTWRVYSTGSNVFQTRAGCLIGAEHVNNS	651
                **************.*********************************************

VM_11-9-21      YECDIPIGAGICASYQTQTNSHRRARSVASQSIIAYTMSLGAENSVAYSNNSIAIPINFT	715
Reference       YECDIPIGAGICASYQTQTNSPRRARSVASQSIIAYTMSLGAENSVAYSNNSIAIPTNFT	719
Q.3             YECDIPIGAGICASYQTQTNSHRRARSVASQSIIAYTMSLGAENSVAYSNNSIAIPINFT	716
VM-9-12-22      YECDIPIGAGICASYQTQTNSHRRARSVASQSIISYTMSLGAENSVAYSNNSIAIPINFT	716
VM_5-1-23       YECDIPIGAGICASYQTQTNSHRRARSVASQSIISYTMSLGAENSVAYSNNSIAIPTNFT	711
                ********************* ************:********************* ***

VM_11-9-21      ISVTTEILPVSMTKTSVDCTMYICGDSTECSNLLLQYGSFCTQLNRALTGIAVEQDKNTQ	775
Reference       ISVTTEILPVSMTKTSVDCTMYICGDSTECSNLLLQYGSFCTQLNRALTGIAVEQDKNTQ	779
Q.3             ISVTTEILPVSMTKTSVDCTMYICGDSTECSNLLLQYGSFCTQLNRALTGIAVEQDKNTQ	776
VM-9-12-22      ISVTTEILPVSMTKTSVDCTMYICGDSTECSNLLLQYGSFCTQLNRALTGIAAEQDKNTQ	776
VM_5-1-23       ISVTTEILPVSMTKTSVDCTMYICGDSTECSNLLLQYGSFCTQLNRALTGIAAEQDKNTQ	771
                ****************************************************.*******

VM_11-9-21      EVFAQVKQIYKTPPIKDFGGFNFSQILPDPSKPSKRSFIEDLLFNKVTLADAGFIKQYGD	835
Reference       EVFAQVKQIYKTPPIKDFGGFNFSQILPDPSKPSKRSFIEDLLFNKVTLADAGFIKQYGD	839
Q.3             EVFAQVKQIYKTPPIKDFGGFNFSQILPDPSKPSKRSFIEDLLFNKVTLADAGFIKQYGD	836
VM-9-12-22      EVFAQVKQIYKTPPIKDFGGFNFSQILPDPSKPSKRSFIEDLLFNKVTFADAGFIKQYGD	836
VM_5-1-23       EVFAQVKQIYKTPPIKDFGGFNFSQILPDPSKPSKRSFIEDLLFNKVTFADAGFIKQYGD	831
                ************************************************:***********

VM_11-9-21      CLGDIAARDLICAQKFNGLTVLPPLLTDEMIAQYTSALLAGTITSGWTFGAGAALQIPFA	895
Reference       CLGDIAARDLICAQKFNGLTVLPPLLTDEMIAQYTSALLAGTITSGWTFGAGAALQIPFA	899
Q.3             CLGDIAARDLICAQKFNGLTVLPPLLTDEMIAQYTSALLAGTITSGWTFGAGAALQIPFA	896
VM-9-12-22      CLGDIAARDLICAQKFNGLTVLPPLLTDEMIAQYTSALLAGTITSGWTFGAGAALQIPFA	896
VM_5-1-23       CLGDIAARDLICAQKFNGLTVLPPLLTDEMIAQYTSALLAGTITSGWTFGAGAALQIPFA	891
                ************************************************************

VM_11-9-21      MQMAYRFNGIGVTQNVLYENQKLIANQFNSAIGKIQDSLSSTASALGKLQDVVNQNAQAL	955
Reference       MQMAYRFNGIGVTQNVLYENQKLIANQFNSAIGKIQDSLSSTASALGKLQDVVNQNAQAL	959
Q.3             MQMAYRFNGIGVTQNVLYENQKLIANQFNSAIGKIQDSLSSTASALGKLQDVVNQNAQAL	956
VM-9-12-22      MQMAYRFNGIGVTQNVLYENQKLIANQFNSAIGKIQDSLSSSASALGKLQDVVNQNAQAL	956
VM_5-1-23       MQMAYRFNGIGVTQNVLYENQKLIANQFNSAIGKIQDSISSSASALGKLQDVVNQNAQAL	951
                **************************************:**:******************

VM_11-9-21      NTLVKQLSSNFGAISSVLNDILARLDKVEAEVQIDRLITGRLQSLQTYVTQQLIRAAEIR	1015
Reference       NTLVKQLSSNFGAISSVLNDILSRLDKVEAEVQIDRLITGRLQSLQTYVTQQLIRAAEIR	1019
Q.3             NTLVKQLSSNFGAISSVLNDILARLDKVEAEVQIDRLITGRLQSLQTYVTQQLIRAAEIR	1016
VM-9-12-22      NTLVKQLSSNFGAISSVLNDILARLDKVEAEVQIDRLITGRLQSLQTYVTQQLIRAAEIR	1016
VM_5-1-23       NTLVKQLSSNFGAISSVLNDILARLDKVEAEVQIDRLITGRLQSLQTYVTQQLIRAAEIR	1011
                **********************:*************************************

VM_11-9-21      ASANLAATKMSECVLGQSKRVDFCGKGYHLMSFPQSAPHGVVFLHVTYVPAQEKNFTTAP	1075
Reference       ASANLAATKMSECVLGQSKRVDFCGKGYHLMSFPQSAPHGVVFLHVTYVPAQEKNFTTAP	1079
Q.3             ASANLAATKMSECVLGQSKRVDFCGKGYHLMSFPQSAPHGVVFLHVTYVPAQEKNFTTAP	1076
VM-9-12-22      ASANLAATKMSECVLGQSKRVDFCGKGYHLMSFPQSAPHGVVFLHVTYVPAQEKNFTTAP	1076
VM_5-1-23       ASANLAATKMSECVLGQSKRVDFCGKGYHLMSFPQSAPHGVVFLHVTYVPAQEKNFTTAP	1071
                ************************************************************

VM_11-9-21      AICHDGKAHFPREGVFVSNGTHWFVTQRNFYEPQIITTDNTFVSGNCDVVIGIVNNTVYD	1135
Reference       AICHDGKAHFPREGVFVSNGTHWFVTQRNFYEPQIITTDNTFVSGNCDVVIGIVNNTVYD	1139
Q.3             AICHDGKAHFPREGVFVSNGTHWFVTQRNFYEPQIITTHNTFVSGNCDVVIGIVNNTVYD	1136
VM-9-12-22      AICHDGKAHFPREGVFVSNGTHWFVTQRNFYEPQTITTHNTFVSGNCDVVIGIVNNTVYD	1136
VM_5-1-23       AICHDGKAHFPREGVFVSNGTHWFVTQRNFYEPQIITTHNTFVSGNCDVVIGIVNNTVYD	1131
                ********************************** ***.*********************

VM_11-9-21      PLQPELDSFKEELDKYFKNHTSPDVDLGDISGINASVVNIQKEIDRLNEVANNLNESLID	1195
Reference       PLQPELDSFKEELDKYFKNHTSPDVDLGDISGINASVVNIQKEIDRLNEVAKNLNESLID	1199
Q.3             PLQPELDSFKEELDKYFKNHTSPDVDLGDISGINASVVNIQKEIDRLNEVAKNLNESLID	1196
VM-9-12-22      PLQPELDSFKEELDKYFKNHTSPNVDLGDIYGINASFVNIQKEIDRLNEVANNLNESLID	1196
VM_5-1-23       PLQPELDSFKEELDKYFKNHTSPDVDLGDISGINASFVNIQKEIDRLNEVANNLNESLID	1191
                ***********************:****** *****.**************:********

VM_11-9-21      LQEFGKYEQYIKWPWYIWLGFIAGLIAIVMVTIMLCCMTSCCSCLKGCCSCGSCCKFDED	1255
Reference       LQELGKYEQYIKWPWYIWLGFIAGLIAIVMVTIMLCCMTSCCSCLKGCCSCGSCCKFDED	1259
Q.3             LQELGKYEQYIKWPWYIWLGFIAGLIAIVMVTIMLCCMTSCCSCLKGCCSCGSCCKFDED	1256
VM-9-12-22      LKELGKYEQYIKWPWYIWLGFIAGLIAIVMVTIMLCCMTSCCSCLKGCCSCGSCCKFDED	1256
VM_5-1-23       LKELGKYEQYIKWPWYIWLGFIAGLIAIVMVTIMLCCMTSCCSCLKGCCSCGSCCKFDED	1251
                *:*:********************************************************

VM_11-9-21      DSEPVLKGVKLHYT	1269
Reference       DSEPVLKGVKLHYT	1273
Q.3             DSEPVLKGVKLHYT	1270
VM-9-12-22      DSEPVLKGVKLHYT	1270
VM_5-1-23       DSEPVLKGVKLHYT	1265
                **************
```
