## Supplemental Figure 2 for "Chronic Shedding of a SARS-CoV-2 Alpha Variant Lineage Q.3/Q.4 in Wastewater"

```
CLUSTAL O(1.2.4) multiple sequence alignment


Reference       MFVFLVLLPLVSSQCVNLTTRTQLPPAYTNSFTRGVYYPDKVFRSSVLHSTQDLFLPFFS	60
VM_11-9-21      MFVFLVLLPLVSSQCVNLTTRTQLPPAYTNSFTRGVYYPDKVFRSSVLHSTQDLFLPFFS	60
VM-9-12-22      MFVFLVLLPLVFSQCVSLTTRTQLPPAYTNSFTRGVYYPDKVFRSSVLYSTQDLFLPFFS	60
CE_2023         MFVFLVLLPLVSSQCVNLTTRTQSPPAYTNSFTRGVYYPDKVFRSSVLYSTQDLFLPFFS	60
VM_5-1-23       MFVFLVLLPLVSSQCVNLKTRTQLTPAYTNSFTRGVYYPDKVFRSSVLYSTQDLFLPFFS	60
                *********** ****.*.****  ***********************:***********

Reference       NVTWFHAIHVSGTNGTKRFDNPVLPFNDGVYFASTEKSNIIRGWIFGTTLDSKTQSLLIV	120
VM_11-9-21      NVTWFHA---SGTNGTKRFDNPVLPFNDGVYFASTEKSNIIRGWIFGTTLDSKTQSLLIV	117
VM-9-12-22      NVTRFQAI--SGTNGIKRFDNPVLPFNDGVYFASTEKSNIIRGWIFGTTLDSKTQSLLIV	118
CE_2023         NVTRFQA---SGTNGIKRFDNPVLPFNDGVYFASTEKSNIIRGWIFGTTLDSKTQSLLIV	117
VM_5-1-23       NVTRFQA---SGTNGIKRFDNPVLPFNDGVYFASTEKSNIIRGWIFGTTLDSKTQSLLIV	117
                *** *:*   ***** ********************************************

Reference       NNATNVVIKVCEFQFCNDPFLGVYYHKNNKSWMESEFRVYSSANNCTFEYVSQPFLMDLE	180
VM_11-9-21      NNATNVVIKVCEFQFCNDPFLG--YHTNNKSWMESEFRVYSSANNCTFEYVSQPFLMDLE	175
VM-9-12-22      NNATNVVIKVCEFQFCNDPFLGV-YHTNNKIWMESEFRVYSSANNCTFEYVSQPFLMDLE	177
CE_2023         NNATNVVIKVCEFQFCNDPFLG--YHTNNKIWMESEFRVYSSANNCTFEYVSQPFLMDLE	175
VM_5-1-23       NNATNVVIKVCEFQFCNDPFLG--YHTNNKIWMESEFRVYSSANNCTFEYVSQPFLMDLE	175
                **********************  **.*** *****************************

Reference       GKQGNFKNLREFVFKNIDGYFKIYSKHTPINLVRDLPQGFSALEPLVDLPIGINITRFQT	240
VM_11-9-21      GKQGNFKNLREFVFKNIDGYFKIYSKHTPINLVRDLPQGFSALEPLVDLPIGINITRFQT	235
VM-9-12-22      EKQGNFKNLREFVFKSIDGYFKIYSKHTPINLVRDLPQGFSALEPLVDLPIGINITRFQT	237
CE_2023         EKQGNFKNLREFVFKSIDGYFKIYSKHTPINLVRDLPQGFSALEPLVDLPIGINITRFQT	235
VM_5-1-23       EKQGNFKNLREFVFKSIDGYFKIYSKHTPINLVRDLPQGFSALEPLVDLPIGINITRFQT	235
                 **************.********************************************

Reference       LLALHRSYLTPGDSSSGWTAGAAAYYVGYLQPRTFLLKYNENGTITDAVDCALDPLSETK	300
VM_11-9-21      LLALHRSYLTPGDSSSGWTAGAAAYYVGYLQPRTFLLKYNENGTITDAVDCALDPLSETK	295
VM-9-12-22      LLALHRSYLTPGDSSSGWTAGAAAYYVGYLQPRTFLLKYNENGTITDAVDCALDPLSETK	297
CE_2023         LLALYRSYLTPGDSSSDWTAGAAAYYVGYLQPRTFLLKYNENGTITDAVDCALDPLSETK	295
VM_5-1-23       LLALHRSSLTPGDSSSDWTAGAAAYYVGYLQPRTFLLKYNENGTITDAVDCALDPLSETK	295
                ****:** ********.*******************************************

Reference       CTLKSFTVEKGIYQTSNFRVQPTESIVRFPNITNLCPFGEVFNATRFASVYAWNRKRISN	360
VM_11-9-21      CTLKSFTVEKGIYQTSNFRVQPTESIVRFPNITNLCPFGEVFNATRFASVYAWNRKRISN	355
VM-9-12-22      CTLKSFTVEKGIYQTSNFRVQPTESIVRFPNITNLCPFGEVFNATRFASVYAWNRKRISN	357
CE_2023         CTLKSFTVEKGIYQTSNFRVQPTESIVRFPNITNLCPFGEVFNATRFASVYAWNRKRISN	355
VM_5-1-23       CTLKSFTVEKGIYQTSNFRVQPTESIVRFPNITNLCPFGEVFNATRFASVYAWNRKRISN	355
                ************************************************************

Reference       CVADYSVLYNSASFSTFKCYGVSPTKLNDLCFTNVYADSFVIRGDEVRQIAPGQTGKIAD	420
VM_11-9-21      CVADYSVLYNSASFSTFKCYGVSPTKLNDLCFTNVYADSFVIRGDEVRQIAPGQTGKIAD	415
VM-9-12-22      CVADYSVLYNSTSFSTFKCYGVSPTKLNDLCFTNVYADSFVIRGDEVRQIAPGQTGKIAD	417
CE_2023         CVADYSVLYNSTSFSTFKCYGVSPTKLNDLCFTNVYADSFVIRGDEVRQIAPGQTGKIAD	415
VM_5-1-23       CVADYSVLYNSTSFSTFKCYGVSPTKLNDLCFTNVYADSFVIKGDEVRQIAPGQTGKIAD	415
                ***********:******************************:*****************

Reference       YNYKLPDDFTGCVIAWNSNNLDSKVGGNYNYLYRLFRKSNLKPFERDISTEIYQAGSTPC	480
VM_11-9-21      YNYKLPDDFTGCVIAWNSNNLDSKVGGNHNYLYRLFRKSNLKPFERDISTEIYQAGSTPC	475
VM-9-12-22      YNYKLPDDFTGCVIAWNSNNLDSKVDGNNNYLFRLFRKSNLKPFERDISTEIYQAGSTPC	477
CE_2023         YNYKLPDDFTGCVIAWNSNNLDS--AGNNNYLFRLFRKSNLKPFERDISTEIYQAGSTPC	473
VM_5-1-23       YNYKLPDDFTGCVIAWNSNNLDS---GNNNYQFRLFRKSKLKPFERDISTEIYQAGNTPC	472
                ***********************   ** ** :******:****************.***


Reference       NGVEGFNCYFPLQSYGFQ-PTNGVGYQPYRVVVLSFELLHAPATVCGPKKSTNLVKNKCV	539
VM_11-9-21      NGVEGFNCYFPLQSYGFRRPTYGVGYQPYRVVVLSFELLHAPATVCGPKKSTNLVKNKCV	535
VM-9-12-22      NGVAGFNCYFPLQSYGFR-PTYGVDHQPYRVVVLSFELLHAPATVCGPKKSTNLVKNKCV	536
CE_2023         NGVVGFNCYFPLKSYGFL-PTYGVDHQPYRVVVLSFELLHAPATVCGPKKSTNLVKNKCV	532
VM_5-1-23       NGVVGFNCYFPLKSYGFL-PTYGVDHQPYRVVVLSFELLHAPATVCGPKKSTNLVKNKCV	531
                *** ********:****  ** **.:**********************************

Reference       NFNFNGLTGTGVLTESNKKFLPFQQFGRDIADTTDAVRDPQTLEILDITPCSFGGVSVIT	599
VM_11-9-21      NFNFNGLTGTGVLTESNKKFLPFQQFGRDIDDTTDAVRDPQTLEILDITPCSFGGVSVIT	595
VM-9-12-22      NFNFNGLTGTGVLTESNKKFLPFQQFGRDIDGTTDAVRDPQTLEILDVTPCSFGGVSVIT	596
CE_2023         NFNFNGLTGTGVLTESNKKFLPFQQFGRDIDGTTDAVRDPQTLEILDVTPCSFGGVSVIT	592
VM_5-1-23       NFNFNGLTGTGVLTESNKKFLPFQQFGRDIDGTTDAVRDPQTLEILDVTPCSFGGVSVIT	591
                ****************************** .***************:************

Reference       PGTNTSNQVAVLYQDVNCTEVPVAIHADQLTPTWRVYSTGSNVFQTRAGCLIGAEHVNNS	659
VM_11-9-21      PGTNTSNQVAVLYQGVNCTEVPVAIHADQLTPTWRVYSTGSNVFQTRAGCLIGAEHVNNS	655
VM-9-12-22      PGTNTSNQVAVLYQGVNCTEVPVAIHADQLTPTWRVYSTGSNVFQTRAGCLIGAEHVNNS	656
CE_2023         PGTNTSNQVAVLYQGVNCTEVPVAIHADQLTPTWRVYSTGSNVFQTRAGCLIGAEHVNNS	652
VM_5-1-23       PGTNTSNQVAVLYQGVNCTEVPVAIHADQLTPTWRVYSTGSNVFQTRAGCLIGAEHVNNS	651
                **************.*********************************************

Reference       YECDIPIGAGICASYQTQTNSPRRARSVASQSIIAYTMSLGAENSVAYSNNSIAIPTNFT	719
VM_11-9-21      YECDIPIGAGICASYQTQTNSHRRARSVASQSIIAYTMSLGAENSVAYSNNSIAIPINFT	715
VM-9-12-22      YECDIPIGAGICASYQTQTNSHRRARSVASQSIISYTMSLGAENSVAYSNNSIAIPINFT	716
CE_2023         YECDIPIGAGICASYQTQTNSHRRARSVASQSIIAYTMSLGAENSVAYSNNSIAIPTNFT	712
VM_5-1-23       YECDIPIGAGICASYQTQTNSHRRARSVASQSIISYTMSLGAENSVAYSNNSIAIPTNFT	711
                ********************* ************:********************* ***

Reference       ISVTTEILPVSMTKTSVDCTMYICGDSTECSNLLLQYGSFCTQLNRALTGIAVEQDKNTQ	779
VM_11-9-21      ISVTTEILPVSMTKTSVDCTMYICGDSTECSNLLLQYGSFCTQLNRALTGIAVEQDKNTQ	775
VM-9-12-22      ISVTTEILPVSMTKTSVDCTMYICGDSTECSNLLLQYGSFCTQLNRALTGIAAEQDKNTQ	776
CE_2023         ISVTTEILPVSMTKTSVDCTMYICGDSTECSNLLLQYGSFCTQLNRALTGIAAEQDKNTQ	772
VM_5-1-23       ISVTTEILPVSMTKTSVDCTMYICGDSTECSNLLLQYGSFCTQLNRALTGIAAEQDKNTQ	771
                ****************************************************.*******

Reference       EVFAQVKQIYKTPPIKDFGGFNFSQILPDPSKPSKRSFIEDLLFNKVTLADAGFIKQYGD	839
VM_11-9-21      EVFAQVKQIYKTPPIKDFGGFNFSQILPDPSKPSKRSFIEDLLFNKVTLADAGFIKQYGD	835
VM-9-12-22      EVFAQVKQIYKTPPIKDFGGFNFSQILPDPSKPSKRSFIEDLLFNKVTFADAGFIKQYGD	836
CE_2023         EVFAQVKQIYKTPPIKDFGGFNFSQILPDPSKPSKRSFIEDLLFNKVTFADAGFIKQYGD	832
VM_5-1-23       EVFAQVKQIYKTPPIKDFGGFNFSQILPDPSKPSKRSFIEDLLFNKVTFADAGFIKQYGD	831
                ************************************************:***********

Reference       CLGDIAARDLICAQKFNGLTVLPPLLTDEMIAQYTSALLAGTITSGWTFGAGAALQIPFA	899
VM_11-9-21      CLGDIAARDLICAQKFNGLTVLPPLLTDEMIAQYTSALLAGTITSGWTFGAGAALQIPFA	895
VM-9-12-22      CLGDIAARDLICAQKFNGLTVLPPLLTDEMIAQYTSALLAGTITSGWTFGAGAALQIPFA	896
CE_2023         CLGDIAARDLICAQKFNGLTVLPPLLTDEMIAQYTSALLAGTITSGWTFGAGAALQIPFA	892
VM_5-1-23       CLGDIAARDLICAQKFNGLTVLPPLLTDEMIAQYTSALLAGTITSGWTFGAGAALQIPFA	891
                ************************************************************

Reference       MQMAYRFNGIGVTQNVLYENQKLIANQFNSAIGKIQDSLSSTASALGKLQDVVNQNAQAL	959
VM_11-9-21      MQMAYRFNGIGVTQNVLYENQKLIANQFNSAIGKIQDSLSSTASALGKLQDVVNQNAQAL	955
VM-9-12-22      MQMAYRFNGIGVTQNVLYENQKLIANQFNSAIGKIQDSLSSSASALGKLQDVVNQNAQAL	956
CE_2023         MQMAYRFNGIGVTQNVLYENQKLIANQFNSAIGKIQDSLSSSASALGKLQDVVNQNAQAL	952
VM_5-1-23       MQMAYRFNGIGVTQNVLYENQKLIANQFNSAIGKIQDSISSSASALGKLQDVVNQNAQAL	951
                **************************************:**:******************

Reference       NTLVKQLSSNFGAISSVLNDILSRLDKVEAEVQIDRLITGRLQSLQTYVTQQLIRAAEIR	1019
VM_11-9-21      NTLVKQLSSNFGAISSVLNDILARLDKVEAEVQIDRLITGRLQSLQTYVTQQLIRAAEIR	1015
VM-9-12-22      NTLVKQLSSNFGAISSVLNDILARLDKVEAEVQIDRLITGRLQSLQTYVTQQLIRAAEIR	1016
CE_2023         NTLVKQLSSNFGAISSVLNDILARLDKVEAEVQIDRLITGRLQSLQTYVTQQLIRAAEIR	1012
VM_5-1-23       NTLVKQLSSNFGAISSVLNDILARLDKVEAEVQIDRLITGRLQSLQTYVTQQLIRAAEIR	1011
                **********************:*************************************

Reference       ASANLAATKMSECVLGQSKRVDFCGKGYHLMSFPQSAPHGVVFLHVTYVPAQEKNFTTAP	1079
VM_11-9-21      ASANLAATKMSECVLGQSKRVDFCGKGYHLMSFPQSAPHGVVFLHVTYVPAQEKNFTTAP	1075
VM-9-12-22      ASANLAATKMSECVLGQSKRVDFCGKGYHLMSFPQSAPHGVVFLHVTYVPAQEKNFTTAP	1076
CE_2023         ASANLAATKMSECVLGQSKRVDFCGKGYHLMSFPQSAPHGVVFLHVTYVPAQEKNFTTAP	1072
VM_5-1-23       ASANLAATKMSECVLGQSKRVDFCGKGYHLMSFPQSAPHGVVFLHVTYVPAQEKNFTTAP	1071
                ************************************************************

Reference       AICHDGKAHFPREGVFVSNGTHWFVTQRNFYEPQIITTDNTFVSGNCDVVIGIVNNTVYD	1139
VM_11-9-21      AICHDGKAHFPREGVFVSNGTHWFVTQRNFYEPQIITTDNTFVSGNCDVVIGIVNNTVYD	1135
VM-9-12-22      AICHDGKAHFPREGVFVSNGTHWFVTQRNFYEPQTITTHNTFVSGNCDVVIGIVNNTVYD	1136
CE_2023         AICHDGKAHFPREGVFVSNGTHWFVTQRNFYEPQIITTHNTFVSGNCDVVIGIVNNTVYD	1132
VM_5-1-23       AICHDGKAHFPREGVFVSNGTHWFVTQRNFYEPQIITTHNTFVSGNCDVVIGIVNNTVYD	1131
                ********************************** ***.*********************

Reference       PLQPELDSFKEELDKYFKNHTSPDVDLGDISGINASVVNIQKEIDRLNEVAKNLNESLID	1199
VM_11-9-21      PLQPELDSFKEELDKYFKNHTSPDVDLGDISGINASVVNIQKEIDRLNEVANNLNESLID	1195
VM-9-12-22      PLQPELDSFKEELDKYFKNHTSPNVDLGDIYGINASFVNIQKEIDRLNEVANNLNESLID	1196
CE_2023         PLQPELDSFKEELDKFFKNHTSPDVDLGDISGINASFVNIQKEIDRLNEVANNLNESLID	1192
VM_5-1-23       PLQPELDSFKEELDKYFKNHTSPDVDLGDISGINASFVNIQKEIDRLNEVANNLNESLID	1191
                ***************:*******:****** *****.**************:********

Reference       LQELGKYEQYIKWPWYIWLGFIAGLIAIVMVTIMLCCMTSCCSCLKGCCSCGSCCKFDED	1259
VM_11-9-21      LQEFGKYEQYIKWPWYIWLGFIAGLIAIVMVTIMLCCMTSCCSCLKGCCSCGSCCKFDED	1255
VM-9-12-22      LKELGKYEQYIKWPWYIWLGFIAGLIAIVMVTIMLCCMTSCCSCLKGCCSCGSCCKFDED	1256
CE_2023         LKELGKYEQYIKWPWYIWLGFIAGLIAIVMVTIMLCCMTSCCSCLKGCCSCGSCCKFDED	1252
VM_5-1-23       LKELGKYEQYIKWPWYIWLGFIAGLIAIVMVTIMLCCMTSCCSCLKGCCSCGSCCKFDED	1251
                *:*:********************************************************

Reference       DSEPVLKGVKLHYT	1273
VM_11-9-21      DSEPVLKGVKLHYT	1269
VM-9-12-22      DSEPVLKGVKLHYT	1270
CE_2023         DSEPVLKGVKLHYT	1266
VM_5-1-23       DSEPVLKGVKLHYT	1265
                **************
```
